# The growth of rural and remote Aboriginal and Torres Strait Islander community laundries: an integrative scoping review

**DOI:** 10.1101/2025.10.16.25337960

**Authors:** K. Summer, D. Nguyen, B. Jones, J. Daw, R. Burgess, R. Wyber

## Abstract

**Objectives and importance of the study:** This article documents rural/remote Aboriginal and Torres Strait Islander community laundries with the aim to support synergistic planning, implementation and evaluation.

**Study type:** An integrative scoping review was conducted in accordance with the Preferred Reporting Items for Systematic Reviews and Meta-Analyses: Scoping Reviews (PRISMA-ScR) guidelines.

**Methods:** The methodology incorporated semi-structured online searches for publicly available grey literature as well as scientific database searches to identify supporting peer-reviewed evidence. Extracted data included: laundry locations; details of establishment, operations and infrastructure; and health and wellbeing impact.

**Results:** At least 55 laundry facilities were established in 38 rural/remote Aboriginal or Torres Strait Islander communities between 2000-2024. Most were established within the past 10 years (*n*=51, 93%) and operated by laundry service providers in partnership with local community organisations (*n*=42, 76%). Laundry locations are publicly available, but we identified no substantiating evidence as to specific health and wellbeing impact.

**Conclusion:** There has been a recent rapid growth in rural/remote Aboriginal and Torres Strait Islander community laundries with plans for future expansion. Equitable access to laundry facilities is tied to human rights to water, sanitation, hygiene and dignity. However, the specific health benefits of community laundries (changes in rates of skin infections, acute rheumatic fever, and rheumatic heart disease) remain unclear. Rigorous evaluations of these health benefits are needed to inform public health policy and community decision making.

## 1. Introduction

The capacity to wash clothing and bedding is the second of nine Healthy Living Practices (HLPs) collectively recognised as determinants of health and wellbeing (Box 1).^1,2^ Codified by the Nganampa Health Council and colleagues in 1987, the HLPs span fundamental human rights, such as those to adequate housing, water, sanitation and dignity.^3,4^ Among benefits to social and emotional health and wellbeing, improvements in living conditions including access to laundry facilities (e.g., hot and cold water, detergents, electricity, washing machines and drying facilities) is linked to the broad prevention of infectious disease. For example, washing clothing and bedding (hereafter referred to as HLP2) is recommended in the management of some skin conditions.^5-8^ Controlling skin pathogens and parasites reduces itch and skin trauma, which can lead to the development of impetigo (skin infection with *Streptococcus pyogenes* [Strep A] and/or *Staphylococcus aureus*) and serious immune-mediated sequelae, including acute rheumatic fever (ARF) and rheumatic heart disease (RHD).^9^ Rural and remote-living Aboriginal and Torres Strait Islander children and young people live with an inequitable burden of impetigo and RHD.^10,11^

### Box 1.

Healthy Living Practices (HLPs), founded on the Nganampa Health’s 1987 Uwankara Palyanyku Kanyintjaku Report (the UPK Report).^1^

- HLP1: Washing people
- **HLP2: Washing clothes and bedding**
- HLP3: Removing wastewater safely
- HLP4: Improving nutrition – including the ability to store, prepare and cook food
- HLP5: Reducing the negative impacts of crowding
- HLP6: Reducing the negative effects of animals, insects and vermin
- HLP7: Reducing the health impacts of dust
- HLP8: Controlling the temperature of the living environment
- HLP9: Reducing minor hazards that cause trauma

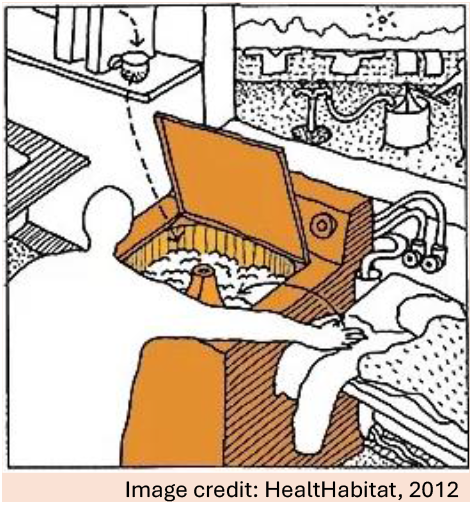

In remote Aboriginal and Torres Strait Islander communities, access to functional household infrastructure and services underpinning access to HLP2 is generally limited..^12-14^ According to recent data from the 2018-2019 National Aboriginal and Torres Strait Islander Health Survey, around 80% of Aboriginal and Torres Strait Islander households in remote/very remote areas have access to working laundry facilities.^15^ However, these figures may be misleading or overestimated: on the basis of surveys of thousands of Aboriginal and Torres Strait Islander households reported in the scientific literature between 2001-2020, access to functional laundry infrastructure and washing machines may be as low as 30%.^16-18^ Statistical data on HLP2 access varies greatly both spatially and temporally. It can reflect access to functional space to install a washing machine but not necessarily a washing machine itself.^16^

The responsibility and impact of variable access to HLP2 are often placed on Aboriginal and Torres Strait Islander households; despite being a consequence of colonisation, geographic and socioeconomic marginalisation, and decades-long policy failure.^13^ Most Aboriginal and Torres Strait Islander families in rural/remote Australia rely on social housing, the standard of which varies greatly between regions.^19^ Basic amenities (a laundry tub and space to install a washing machine) should be provided^20^, but washing machines are not.^21,22^ Barriers to accessing HLP2 at a household level relate to washing machine procurement (suitability, price, availability, transport), heavy machine usage and wear (due to overcrowding, demand sharing, and deterioration of mechanical parts by hard water and dust), cultural factors (avoidance rules), and constrained access to adequate housing, water, electricity, consumables, maintenance, and repairs.^22-24^

Publicly accessible community laundries with industrial-scale machine washers and dryers have long been identified as a mechanism to improve access to HLP2 in remote Aboriginal and Torres Strait Islander communities.^23,25^ Two leading organisations, Aboriginal Investment Group (AIG) and Orange Sky, have launched specific remote community laundry programs of work in recent years,^26,27^ whilst the Heart Foundation 2024/25 Federal Budget Submission called for Commonwealth investment in 70 remote community laundries over the next five years.^28^ Advocates argue for the potential of community laundries to improve a range of health and wellbeing outcomes, with strong emphasis on reducing the burden of skin infections, ARF and RHD.^28-30^

There has not yet been a structured documentation of community laundries to facilitate a unified approach to planning and evaluation. Nor is there a synthesis of published academic literature/data demonstrating their impact in these settings, although some impact frameworks have been commissioned.^31-33^ These unknowns represent gaps, and possible missed opportunities, to support access to HLP2, and better understand and promote the specific health benefits^34^ of community laundry services.

The aim of this work was to document community laundries located in rural and remote Aboriginal and Torres Strait Islander communities in Australia. Characterising the locations and growth of these facilities is an important foundational step for considering synergistic planning and equity of access, exploring effective models of delivery, and enabling monitoring and evaluation of health and wellbeing impact. This review is timely and important given the recent focus on community laundry facilities and advocacy for further investment and expansion.^35,36^

## 2. Methods

### 2.1 Identification of community laundry facilities

We conducted an integrative, systematic scoping review following the Preferred Reporting Items for Systematic reviews and Meta-Analyses extension for Scoping Reviews (PRISMA-ScR) guidelines.^37^ The methodology was chosen to identify community laundry locations documented in grey literature (websites, media releases, community newsletters, technical reports) as well as in peer-reviewed publications (should a community laundry have been documented, studied or evaluated academically). In the absence of any other unified source of information, and in light of urgent policy need, this approach enabled us to understand the locations of community laundry facilities (across all providers and models), estimate rates of growth, and identify any supporting scholarly information (or lack thereof) as to health and wellbeing impact.

Online databases Scopus, PubMed and Informit were searched using terms relevant to “Aboriginal and Torres Strait Islander community” and “community laundry” in the Australian context for literature published from 2000 to 2024 (as of November 2024). A full description of the search strategy is available in Supplementary Material. Since relevant peer-reviewed publications were scant, we pragmatically sought publicly available information by conducting structured online searches. Grey literature searches were firstly conducted within websites of key remote community laundry service providers (Orange Sky and AIG) (Figure 1). This was supplemented by structured searches in Google using terms relevant to “Aboriginal and Torres Strait Islander community” and “community laundry” plus iterative combinations of community names listed on the Australian Government National Indigenous Australians Agency (NIAA) website^38^ (Figure 1). This aligned with an academic database search methodology, for example, [“Aboriginal and Torres Strait Islander” OR “Indigenous” OR “community” OR “<specific NIAA listed community name>“] AND [“laundry” OR “washing machine”]. Only the first five pages of Google results were considered relevant.

**Figure 1.**
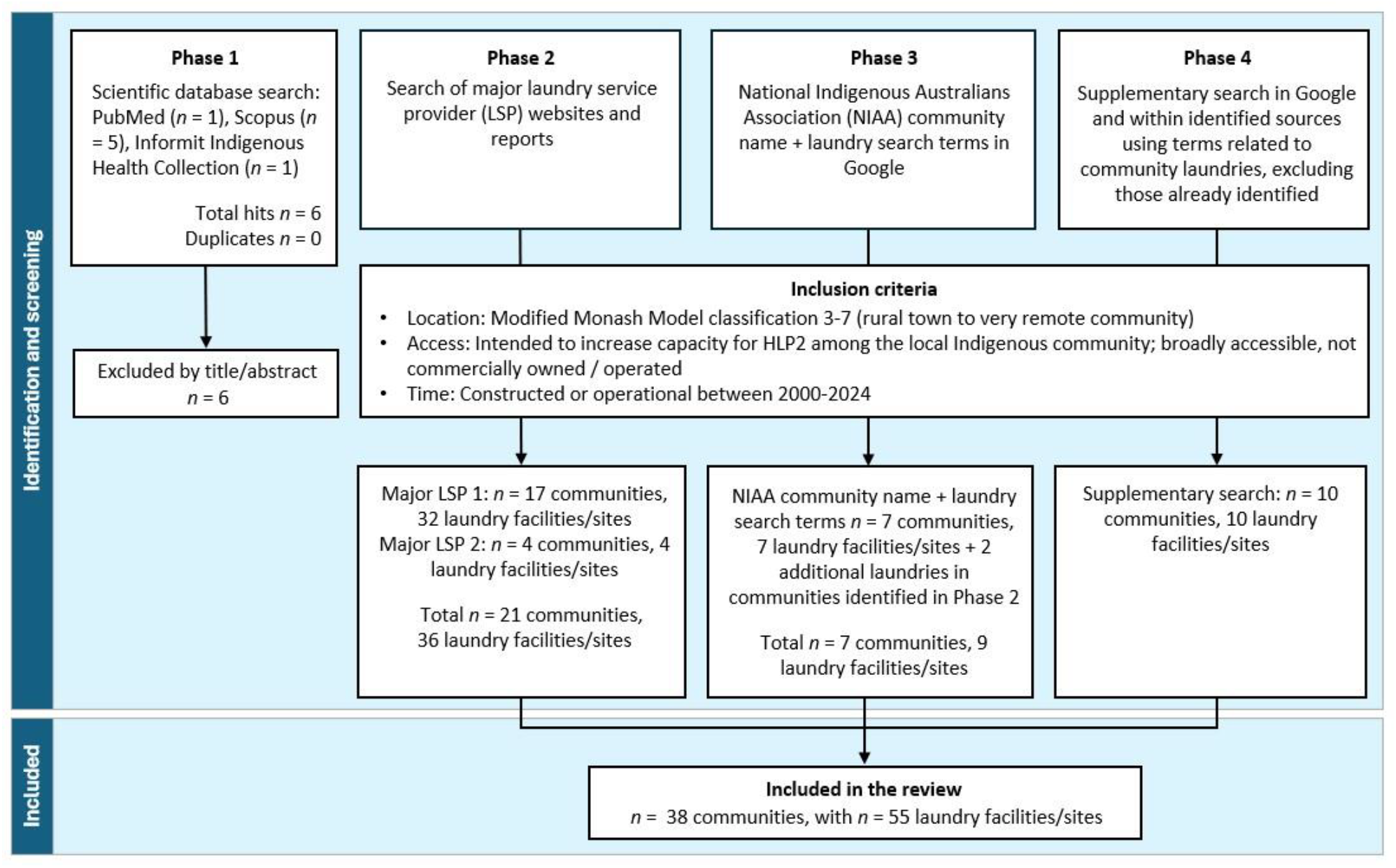
Flow chart showing the process of identification of Aboriginal and Torres Strait Islander community laundries for inclusion in this review.

Laundries were considered relevant for inclusion in the review if they were: located in rural or remote areas of Australia according to the Modified Monash Model (MMM) ^39^ (classifications 3-7); a public asset (not privately/commercially owned-operated laundromats); intended to increase access to HLP2 among the local Aboriginal or Torres Strait Islander community by way of initiation and/or partnership with a local Aboriginal or Torres Strait Islander community controlled organisation; and constructed, funded or temporarily operational during or after the year 2000 (Figure 1). Very transient laundry services (e.g., mobile services present in a community for less than one month in response to a natural disaster or to support an event) were excluded since they were considered unique models/circumstances. Only laundries present for *at least* one month were considered likely to require significant funding and ongoing supports, and impact health and wellbeing. New laundries earmarked for establishment in specific communities within the next 12 months were noted. Large scale residential laundry initiatives (i.e., supply of washing machines to individual households) were noted but not included in the analysis. The presence of single washing machines located in schools, health clinics and arts centres were unable to be reliably identified but may be widely used. These inclusion criteria were workshopped repeatedly until the appropriate working definition was reached.

### 2.2 Data synthesis and analysis

Data were collated in Excel using an agreed data extraction template capturing details regarding laundry locations (including jurisdiction/region, classification of remoteness [MMM], and community population size serviced by the laundry), establishment (provider and partnerships, type of service e.g., mobile or fixed structure), operations (e.g., running costs and access to consumables), and health and wellbeing impact or evaluation. All data were independently extracted and checked by two reviewers (KS and DN). Any discrepancies were resolved by RW and RB. Descriptive statistics were calculated in Excel. The total number of rural/remote community laundries in each jurisdiction were overlaid on a map originally sourced from the NIAA website (https://www.niaa.gov.au/niaa-corporate-plan-2023-24/our-operating-context) and adapted using Canva. Although the locations of all included community laundries are publicly available, specific community names are not disclosed in this manuscript. We focused on access to HLP2 by assigning broad categories of establishment and operation rather than referring to specific laundry service providers.^1^

### 2.3 Quality assessment and conduct

Sources of data included in this review were not eligible for screening against a formal quality assessment tool. Grey literature sources were included but acknowledged as being of generally low-quality evidence. This review was co-designed and co-authored by First Nations and non-First Nations collaborators, in response to research priorities established by the Indigenous Governance Council (IGC) for a National Health and Medical Research Council (NHMRC)-funded program of work. Conduct and reporting were guided and reported against each item in the CONSIDER statement^40^ (Supplementary Material).

## 3. Results

### 3.1 Identification of community laundry facilities

Figure 1 summarises the search process. Scientific database searches (Phase 1) returned six results but zero relevant articles. Targeted and snowball searches in Google (Phases 2-4) identified 38 rural or remote communities with 55 public laundry facilities/sites established between 2000-2024 intended to increase capacity for HLP2 among local Aboriginal and Torres Strait Islander people (Figure 1). Expanded data and references are supplied in Supplementary Material.

### 3.2 Establishment and operation

The 55 identified community laundries were recently established in 38 communities predominantly located in the Northern Territory (*n*=21, 55%), Western Australia (*n*=9, 24%), and northern Queensland (*n*=7, 18%); there was only one community with a laundry located in South Australia and none in New South Wales (NSW) or southern Queensland (Table 1; Figure 2). Most laundries were in communities classified as either remote (MMM 6) or very remote (MMM 7) locations (95%, *n*=36), with the remaining located in rural towns (MMM5) (Table 1; Figure 2). In some places, mobile services were delivered at several fixed locations (sites) within the same community (Table 1). Additionally, we noted at least fourteen additional remote communities earmarked to receive laundries in the near future (Figure 3).^26,27,31,36,41^

**Table 1.**
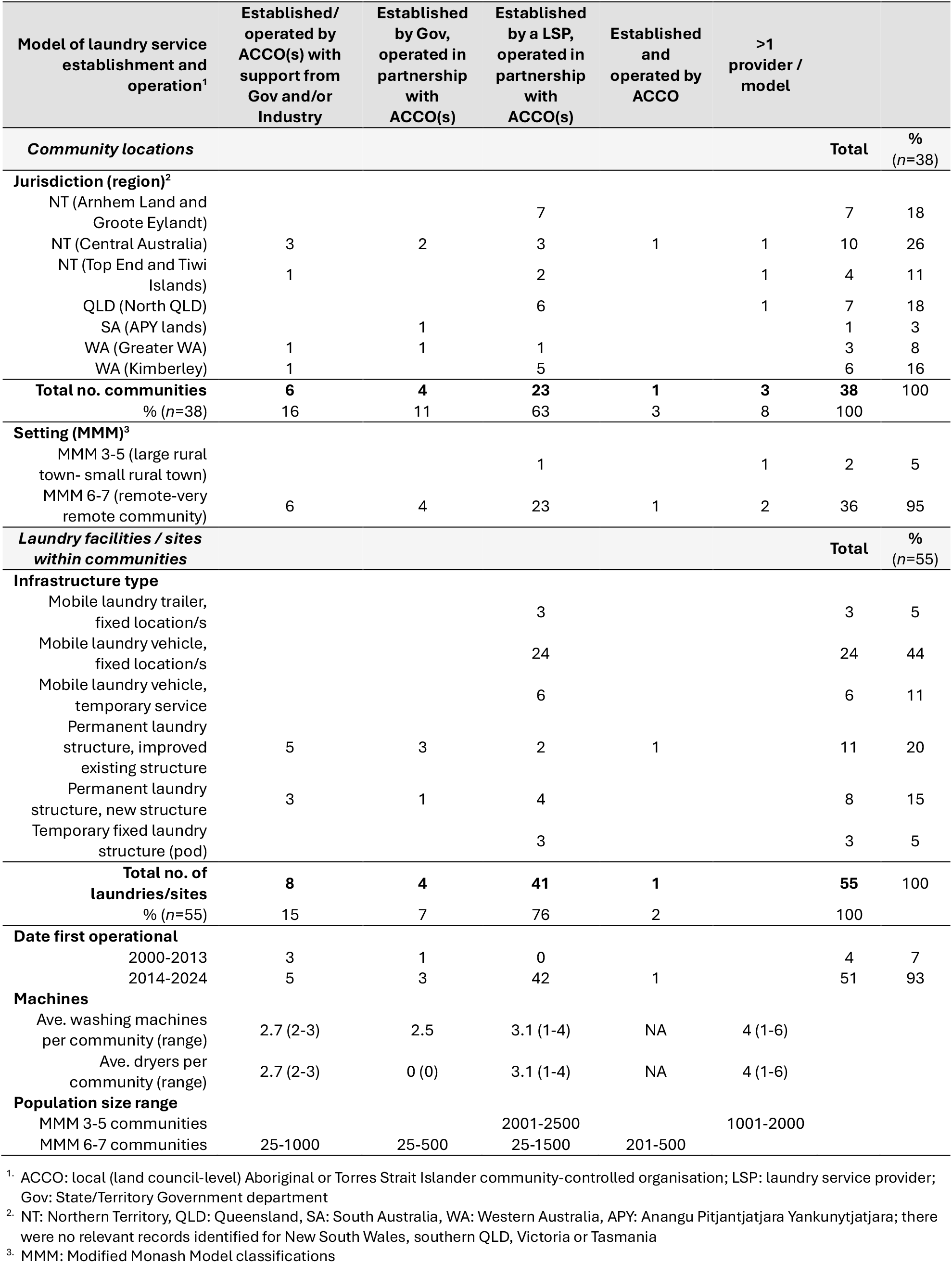
Summary of community laundries established in rural and remote Aboriginal and Torres Strait Islander communities between 2000-2024 identified in a scoping review of publicly available information.

| Model of laundry service establishment and operation<sup>1</sup> | Established/operated by ACCO(s) with support from Gov and/or Industry | Established by Gov, operated in partnership with ACCO(s) | Established by a LSP, operated in partnership with ACCO(s) | Established and operated by ACCO | >1 provider / model |  |  |
| --- | --- | --- | --- | --- | --- | --- | --- |
| Community locations |  |  |  |  |  | Total | %<br>(n=38) |
| **Jurisdiction (region)<sup>2</sup>** |  |  |  |  |  |  |  |
| NT (Arnhem Land and Groote Eylandt) |  |  | 7 |  |  | 7 | 18 |
| NT (Central Australia) | 3 | 2 | 3 | 1 | 1 | 10 | 26 |
| NT (Top End and Tiwi Islands) | 1 |  | 2 |  | 1 | 4 | 11 |
| QLD (North QLD) |  |  | 6 |  | 1 | 7 | 18 |
| SA (APY lands) |  | 1 |  |  |  | 1 | 3 |
| WA (Greater WA) | 1 | 1 | 1 |  |  | 3 | 8 |
| WA (Kimberley) | 1 |  | 5 |  |  | 6 | 16 |
| **Total no. communities** | **6** | **4** | **23** | **1** | **3** | **38** | **100** |
| % (n=38) | 16 | 11 | 63 | 3 | 8 | 100 |  |
| **Setting (MMM)<sup>3</sup>** |  |  |  |  |  |  |  |
| MMM 3-5 (large rural town- small rural town) |  |  | 1 |  | 1 | 2 | 5 |
| MMM 6-7 (remote-very remote community) | 6 | 4 | 23 | 1 | 2 | 36 | 95 |
| **Laundry facilities / sites within communities** |  |  |  |  |  | Total | %<br>(n=55) |
| **Infrastructure type** |  |  |  |  |  |  |  |
| Mobile laundry trailer, fixed location/s |  |  | 3 |  |  | 3 | 5 |
| Mobile laundry vehicle, fixed location/s |  |  | 24 |  |  | 24 | 44 |
| Mobile laundry vehicle, temporary service |  |  | 6 |  |  | 6 | 11 |
| Permanent laundry structure, improved existing structure | 5 | 3 | 2 | 1 |  | 11 | 20 |
| Permanent laundry structure, new structure | 3 | 1 | 4 |  |  | 8 | 15 |
| Temporary fixed laundry structure (pod) |  |  | 3 |  |  | 3 | 5 |
| **Total no. of laundries/sites** | **8** | **4** | **41** | **1** |  | **55** | **100** |
| % (n=55) | 15 | 7 | 76 | 2 |  | 100 |  |
| **Date first operational** |  |  |  |  |  |  |  |
| 2000-2013 | 3 | 1 | 0 |  |  | 4 | 7 |
| 2014-2024 | 5 | 3 | 42 | 1 |  | 51 | 93 |
| **Machines** |  |  |  |  |  |  |  |
| Ave. washing machines per community (range) | 2.7 (2-3) | 2.5 | 3.1 (1-4) | NA | 4 (1-6) |  |  |
| Ave. dryers per community (range) | 2.7 (2-3) | 0 (0) | 3.1 (1-4) | NA | 4 (1-6) |  |  |
| **Population size range** |  |  |  |  |  |  |  |
| MMM 3-5 communities |  |  | 2001-2500 |  | 1001-2000 |  |  |
| MMM 6-7 communities | 25-1000 | 25-500 | 25-1500 | 201-500 |  |  |  |

<sup>1</sup>. ACCO: local (land council-level) Aboriginal or Torres Strait Islander community-controlled organisation; LSP: laundry service provider; Gov: State/Territory Government department
<sup>2</sup>. NT: Northern Territory, QLD: Queensland, SA: South Australia, WA: Western Australia, APY: Anangu Pitjantjatjara Yankunytjatjara; there were no relevant records identified for New South Wales, southern QLD, Victoria or Tasmania
<sup>3</sup>. MMM: Modified Monash Model classifications

**Figure 2.**
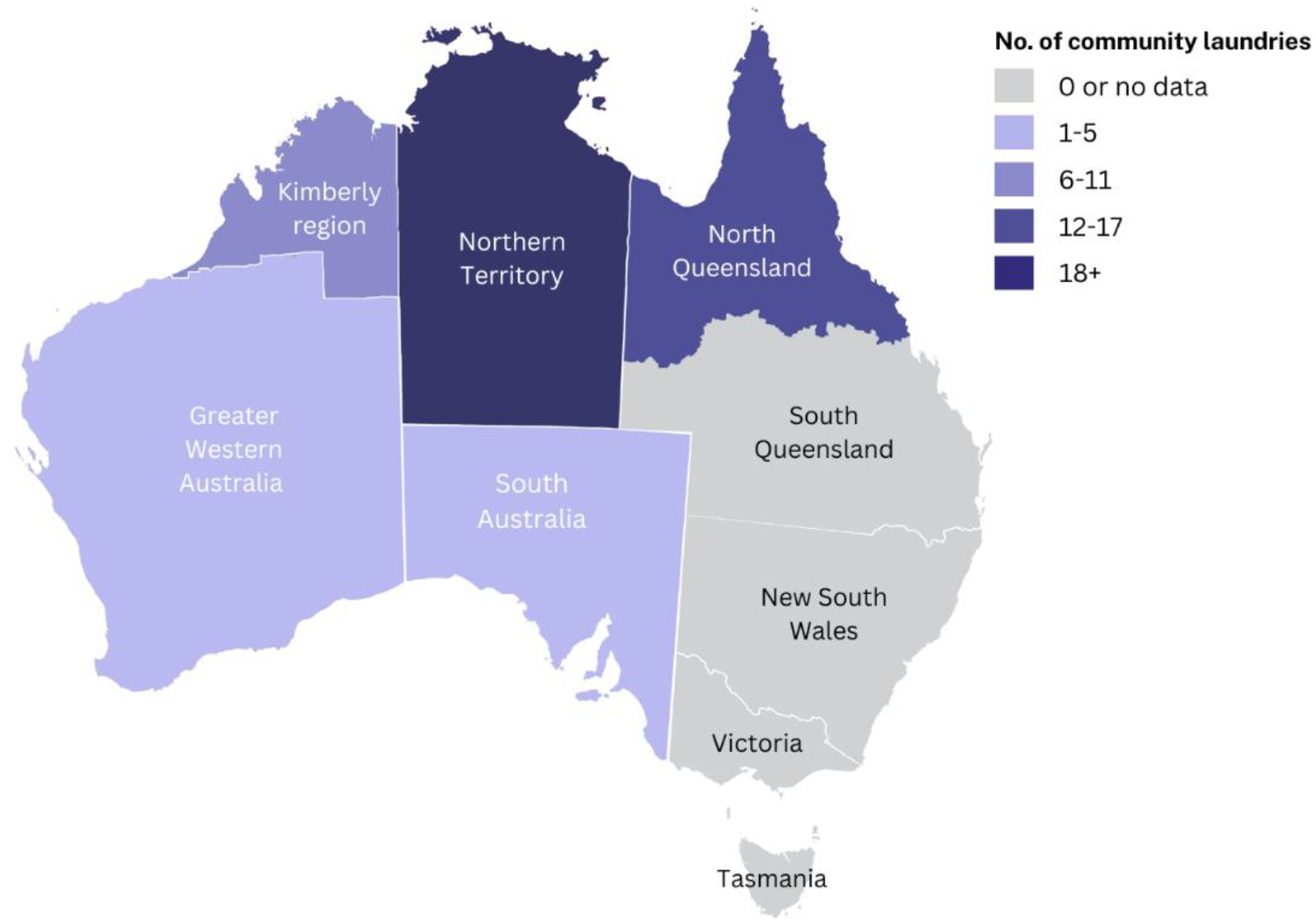
Map of Australia separated by jurisdiction/region with colour intensity representing the density of community laundries established between 2000-2024 intended to increase access to HLP2 for rural and remote-living Aboriginal and Torres Strait Islander people.

**Figure 3.**
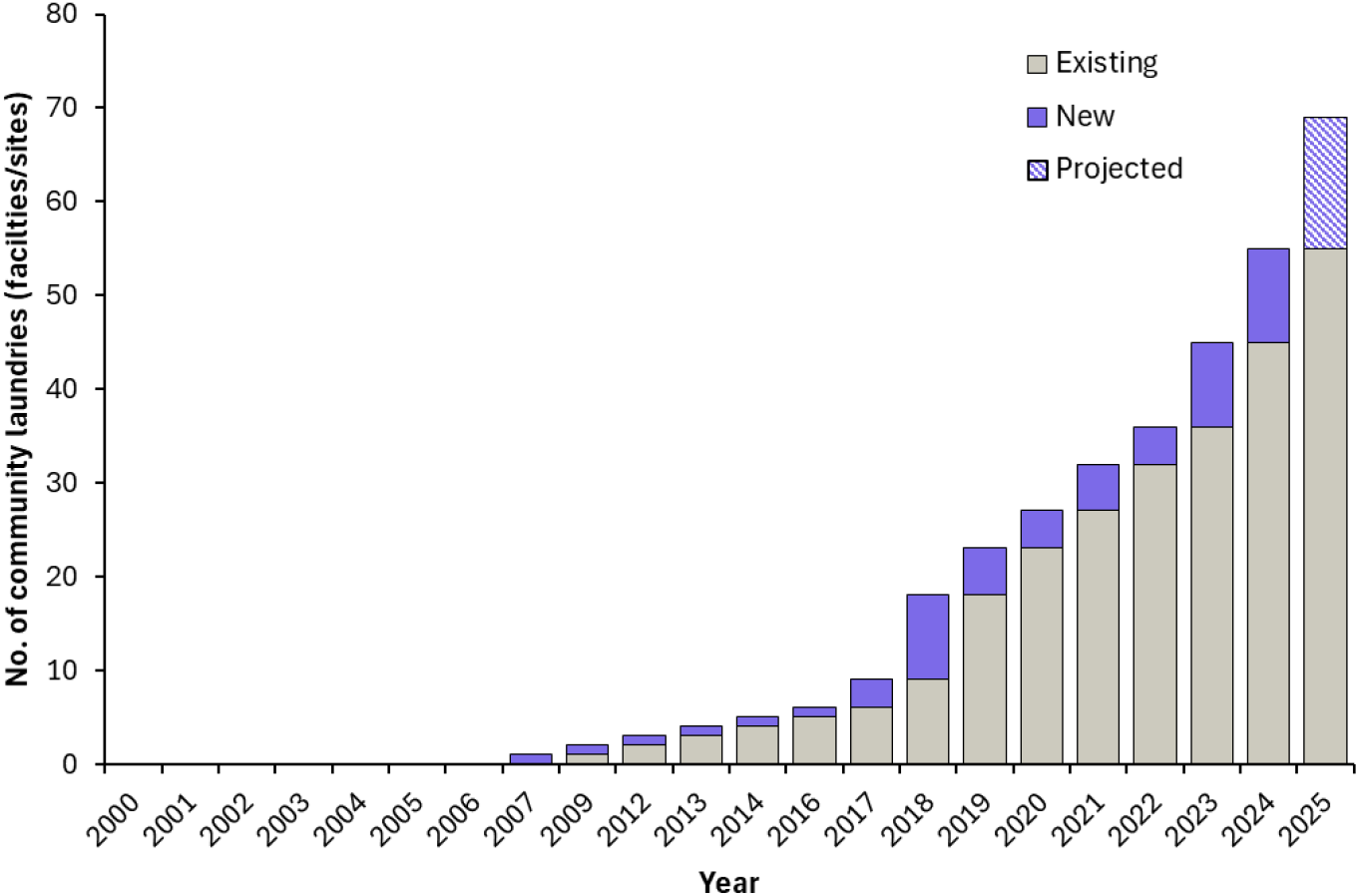
Cumulative growth of community laundries identified in this review established in rural and remote Aboriginal and Torres Strait Islander communities between the years 2000-2024. Projections for 2025 are based on laundry service provider reports and websites. Community laundries may have been established prior to 2000 but were outside of the scope of this work. Additional laundry facilities may have been established or proposed since the time this review was conducted (Nov, 2024).

There were a range of different models by which the 55 identified laundries were established and operated. Eight laundries (15%) were established or initiated by a local Aboriginal or Torres Strait Islander community organisation with support from respective State/Territory governments or an industry partner, usually in the form of a grant (Table 1). Less frequently (*n*=4 laundries, 7%), the establishment of a community laundry was driven by a State/Territory government department and subsequently operated by a local Aboriginal or Torres Strait Islander community organisation (Table 1). Only one laundry appeared to have been established and operated by a local land council-level Aboriginal or Torres Strait Islander community organisation without an additional partnering organisation (Table 1). In 76% of cases (*n*=42 laundries), the laundry was established by a laundry service provider and operated in partnership with local Aboriginal or Torres Strait Islander community organisation(s) (Table 1). In three communities, there was more than one provider or type of laundry (e.g., permanent structure and mobile service) (Table 1).

Four community laundries (7%) were constructed between 2007-2013; these were all established/operated in partnership between local community organisations and State/Territory governments and often involved retrofitting an existing community laundry building or adding a laundry facility to an existing service, such as a women’s centre (Table 1; Figure 3). There were no new laundries established between 2000-2006 (Figure 3). The growth of laundry service provider-established community laundries began in 2017 (Figure 3). Fifty-one laundries (93%) were established within the past 10 years (Table 1), and of these 31 (56%) were established within the past 5 years (2020-2024) (Table 1; Figure 3). These more recent models were largely provided as new permanent laundry structures, mobile trucks and trailers (either temporary or scheduled to appear at fixed locations), or semi-permanent structures/transportable pods (Table 1). Altogether, 27 community laundries (49%) were operated as scheduled mobile services appearing at fixed sites, whilst 19 (35%) were permanent structures, and 9 (16%) were provided as mobile or semi-permanent structures that were temporary but present in a community for over a month at a given time (Table 1).

Detailed information on equipment (such as machine technical specifications, clothes lines, and tubs), access to consumables, and costs were often unavailable. Laundry facilities always included dryers when a laundry service provider was involved but were less common under other arrangements (Table 1). Laundry service providers also usually supplied detergents, though the formulation, brands, and other details of these products were unspecified. Detergents used in community laundries established without a laundry service provider were not mentioned.

Estimated local Aboriginal and Torres Strait Islander populations ranged from 25 to 1,000 people in remote/very remote communities, and between 1,000-2,500 people in rural communities (Table 1); the ratio of people to washing machines/dryers where an average laundry facility provides three washing machines and three dryers is therefore approximately 1:8-300 in remote/very remote communities and 1:300-800 in rural towns. Based on scant publicly available financial data (six records only), the cost of establishing a rural/remote community laundry facility can range from $AUD 74,000 to 406,863, with an average cost of $AUD 235,000 (Supplementary Material).

There was a paucity of independent, peer-reviewed published evidence relating to the health and wellbeing impacts of the identified community laundries for rural and remote-living Aboriginal and Torres Strait Islander people (Figure 1). Nonetheless, promotion and health messaging surrounding community laundries focused almost exclusively on their benefits and positive impacts. This included anecdotal claims regarding physical health (e.g., significant reductions in rates of skin infections, scabies, ARF, RHD, and trachoma; improved health outcomes), hygiene (e.g., improved hygiene, understanding of hygiene, high hygienic wash standards; ensured access to essential hygiene infrastructure; promoting clean and healthy living), and social and emotional health and wellbeing (e.g., improved sleep, wellbeing and quality of life; a hub of conversation and education; increased local employment and capacity building; meaningful outcomes; solving problems; aligning with health, social and cultural community needs; creating resilient and strong communities) (Supplementary Material). There were no publicly available references to unintended consequences or potential risks associated with specific community laundries.

## 4. Discussion

This review demonstrates a rapid acceleration in the establishment of rural and remote Aboriginal and Torres Strait Islander community laundries since the year 2017, with significant growth in the past five years. These laundries, which vary by design as permanent/semi-permanent structures or mobile trucks/trailers, are unified in having been predominantly established by laundry service providers and operated in partnership with local Aboriginal or Torres Strait Islander people and community-controlled organisations, with the intention to increase access to HLP2 and improve health and wellbeing outcomes. Locations of community laundries and broad public health claims were identifiable in publicly available online grey literature, but independent substantiating evidence as to their actual impact on health and wellbeing was not found, either in publicly available online sources or peer-reviewed literature.

The value of biomedical evidence in decision-making about social determinants of health, and human rights is complex. Western methodologies which centre reductionist approaches to the production and interpretation of biomedical evidence are not always meaningful or appropriate in Aboriginal and Torres Strait Islander health and social settings.^42^ Throughout the 20^th^ century, broad improvements in health, wellbeing, and life expectancy were achieved in Australia and internationally; raised standards of living, along with health promotion and medical advances, contributed to this gain.^43^ There are justified reasons therefore to assume that increasing access to HLP2, as one means to improve living conditions, will broadly improve health and wellbeing in rural and remote Aboriginal and Torres Strait Islander communities.

However, specific health outcomes causally related to particular interventions should be evidence informed.^34,44^ Ethical health promotion involves providing accurate information and advice that enables individuals, communities, and governments to make active choices about how to invest time, energy, and resources to achieve specific outcomes.^34^ This means that statements about specific health impacts, including changes in rates of skin infections and infestations (e.g., impetigo and scabies, respectively), and post-streptococcal infection sequelae (such as ARF and RHD) associated with the introduction of Aboriginal and Torres Strait Islander community laundries should be based on rigorous evaluation.

In order to eliminate pathogens and parasites from fabric fomites and thus reduce transmission of skin infections and infestations, it is likely that thresholds for effective laundering must be reached. For example, Bernigaud *et al*. (2020) established the thermal killing point for scabies mites and eggs as ≥50°C for at least 10 min achievable by machine washing or drying.^6^ Similarly, exposure to hot washing and/or drying (>60°C) for at least 15 min has been shown to be effective against bed bugs, fungal pathogens, *S. aureus*, and head lice.^45-49^ It is unclear whether these thresholds for infection control and the relevant Australian standard for laundry practice (AS 4146:2024)^50^ are applied or adhered to in real world community contexts. Community laundries may also pose unique infection risks to be managed.^51,52^ Some organisms, such as *S. aureus*, bed bugs, dermatophytes, are known to be transmitted via fabric fomites, whilst evidence for fabric fomite transmission of scabies, head lice, and Strep A is very limited.^53^ A review of the role of fabrics in the transmission of skin pathogens/ectoparasites, and technical specifications for laundering to control them, would provide useful guidance and is currently underway (Prospero registration: CRD42024594116).^53^

Parallels can be drawn between the strong advocacy for community laundries in recent years and that for community swimming pools throughout the 1990’s and early 2000’s.^54,55^ During that time, the benefits of swimming pools for rural and remote Aboriginal and Torres Strait Islander communities were actively promoted, including reduced rates of skin, ear and eye infections, improved economic participation, and enhanced social and emotional wellbeing.^54,56^ In 2016, Hendrickx *et al*.^57^ undertook a systematic review of 12 studies investigating health outcomes associated with swimming pools, concluding that there was a consistent decline in the prevalence and severity of skin sores associated with a new pool opening or implementation of a community-based swimming program. However, evidence around ear and eye infections were inconclusive, and the social and emotional wellbeing benefits of community swimming pools remained conjectural or anecdotal.^57,58^ The authors also noted that the potential risks associated with swimming pools (e.g., water safety) were rarely discussed, and that confounding effects of other ongoing programs or public health interventions within selected communities were not controlled for. Because the magnitude and sustainability of positive impacts arising from community pools depend on patterns of use, costs, governance and sustainability,^57,59^ there is now more guarded optimism around community swimming pools.^59,60^ Lessons learned from past enthusiasm for, and insufficient planning and evaluation of, community swimming pools can be applied to community laundries. Both forms of infrastructure have commonly accepted health and wellbeing benefits, in addition to high infrastructure costs, potential risks, maintenance challenges, and available alternatives.

Large scale rollout of community laundries evidently has, and will continue to, necessitate significant investment of financial and social capital. Based on average costs calculated using available financial data, assets established since the year 2000 may be valued at around $AUD 13 million. Additionally, annual ongoing operation and maintenance costs (sometimes including employment costs for local staff) have been estimated at over $100,000 per laundry^31^ (around $AUD 5.5 million/year for current laundries). At a conservative estimate, the cost of an additional 100 laundries over ten years could exceed $AUD 120 million. Funding committed on the basis of specific health benefits should therefore be supported by mechanisms for quality assurance and evaluation.

Improvement of housing and living conditions for Aboriginal and Torres Strait Islander people is a highly politicised and contested space. Whilst community laundries present opportunities for potential health benefits, strategic resourcing, and genuine partnerships, they also hold potential to inadvertently compete with funding for housing and health services, which is being urgently called for by Aboriginal and Torres Strait Islander peak organisations.^61,62^

There are several acknowledged limitations to this review. Firstly, relying on publicly available data may have missed or misrepresented some community laundries, laundry service providers or community partnerships. We did not validate online reports or seek direct input from key stakeholders on this manuscript, as a means to maintain an objective methodology. Secondly, we developed an operational definition of rural/remote Aboriginal or Torres Strait Islander community laundries for inclusion/exclusion purposes which may not reflect community or contextual understanding. Thirdly, we cannot be confident that all included laundries continue to be operational; it is likely that openings of laundries have received more media coverage than closures, leading to potential bias. Nonetheless, in the absence of any other unified source of information, this document may inform critical discussion and decision making.

## 5. Conclusion

Community laundries are increasingly heralded as a means to increase access to HLP2 and thereby address health and wellbeing inequities for rural and remote-living Aboriginal and Torres Strait Islander people. The number of rural/remote Aboriginal and Torres Strait Islander community laundries has increased rapidly in recent years, and more are in the pipeline. Access to HLP2 is closely tied to human rights and broad improvements standards of living. However, promotion of specific health benefits causally associated with community laundries (e.g., reduction in rates of skin infections, ARF, and RHD) should be evidence based.

## Acknowledgements

We acknowledge and sincerely thank the Indigenous Governance Council for the STARFISH project for their leadership, in addition to the wider STARFISH Investigator team and Ms. Ainslie Poore.

## Credit authorship statement

KS: methodology, investigation, formal analysis, writing – original draft. DN: methodology, investigation. BJ: supervision, data interpretation, writing – review and editing. JD: investigation, writing – review and editing. RB: supervision, writing – review and editing. RW: conceptualisation, methodology, supervision, writing – review and editing.

## Authorship inclusivity and diversity statement

One or more of the manuscript authors self-identifies as being of Aboriginal or Torres Strait Islander origin.

## Data availability statement

All data generated/analysed during this study are provided as Supplementary Material. Additional data are available on request of the corresponding author.

## Footnotes

1 There are a variety of organisational and governance structures among laundry service providers, including non-government organisations (NGO’s), Aboriginal-owned organisations, and partnerships with other groups. The specifics of these arrangements are outside the scope of this review.

## References

1. Nganampa Health Council. Report of Uwankara Palyanyku Kanyintjaku: an environmental and public health review within the Anangu Pitjantjatjara Lands. South Australia: Nganampa Health Council Inc, South Australian Health Commission, Aboriginal Health Organisation of South Australia; 1987. pp. 101

2. Australian Government Department of Families, Housing, Community Services, and Indigenous Affairs (DFHCSIA). National Indigenous housing guide: improving the living environment for safety, health and sustainability (3^rd^ ed.). Canberra: DFHCSIA; 2007. pp. 16

3. Office of the High Commissioner for Human Rights (OHCHR). OHCHR and the rights to water and sanitation. OCHR; 2024 [cited Nov, 2024]. Available from: https://www.ohchr.org/en/water-and-sanitation

4. Australian Government Attorney-General’s Department (AGD). Right to an adequate standard of living, including food, water and housing. AGD; 2025 [cited August, 2025]. Available from: https://www.ag.gov.au/rights-and-protections/human-rights-and-anti-discrimination/human-rights-scrutiny/public-sector-guidance-sheets/right-adequate-standard-living-including-food-water-and-housing

5. Balge MZ, Krieger GR. Lice, mites, bedbugs, fleas, tumbu flies: disease-causing agents preventable through appropriate laundry practices, sanitation regimes, and hygiene. Paper presented at the SPE International Health, Safety and Environment Conference, Abu Dhabi, UAE; April 2006. SPE-98866-MS

6. Bernigaud C, Fernando DD, Lu H et al. How to eliminate scabies parasites from fomites: a high-throughput ex vivo experimental study. J Am Acad Dermatol 2020; 83(1):241–245.

7. Bloomfield SF, Exner M, Signorelli C et al. The infection risks associated with clothing and household linens in home and everyday life settings, and the role of laundry. Montacute, United Kingdon: International Scientific Forum on Home Hygiene; 2011. pp. 47 [cited Nov, 2024]. Available: https://ifh-homehygiene.org/review-best-practice/infection-risks-associated-clothing-and-household-linens-home-and-everyday-life/

8. The Australian Healthy Skin Consortium. National healthy skin guideline: for the diagnosis, treatment and prevention of skin infections for Aboriginal and Torres Strait Islander children and communities in Australia. Perth: Telethon Kids Institute; 2023. pp. 132 [cited Nov, 2024]. Available: https://infectiousdiseases.thekids.org.au/resources/skin-guidelines/

9. McDonald MI, Towers RJ, Andrews RM et al. Low rates of streptococcal pharyngitis and high rates of pyoderma in Australian Aboriginal communities where acute rheumatic fever is hyperendemic. Clin Infect Dis 2006; 43(6):683–689.

10. Wiegele S, McKinnon E, van Schaijik B et al. The epidemiology of superficial Streptococcal A (impetigo and pharyngitis) infections in Australia: a systematic review. PLoS One 2023; 18(11):e0288016.

11. Wyber R, Wade V, Anderson A et al. Rheumatic heart disease in Indigenous young peoples. Lancet Child Adolesc Health 2021; 5(6):437–446.

12. Bailie RS, Wayte KJ. Housing and health in Indigenous communities: key issues for housing and health improvement in remote Aboriginal and Torres Strait Islander communities. Aust J Rural Health 2006; 14(5):178–183.

13. Habibis D, Phillips R, Phibbs P. Housing policy in remote Indigenous communities: how politics obstructs good policy. Housing Studies 2019; 34(2):252–271.

14. Memmott P, Lansbury N, Go-Sam C et al. Aboriginal social housing in remote Australia: crowded, unrepaired and raising the risk of infectious diseases. Global Discourse 2022; 12(2):255–284.

15. Australian Institute of Health and Welfare (AIHW). Determinants of health: 2.02 Access to functional housing with utilities. AIHW; 2024 [cited Nov, 2024]. Available from: https://www.indigenoushpf.gov.au/measures/2-02-access-to-functional-housing-with-utilities

16. Standen JC, Morgan GG, Sowerbutts T et al. Prioritising housing maintenance to improve health in Indigenous communities in NSW over 20 years. Int J Env Health Res 2020; 17(16).

17. Torzillo PJ, Pholeros P, Rainow S et al. The state of health hardware in Aboriginal communities in rural and remote Australia. Aust N Z J Public Health 2008; 32(1):7–11.

18. Bailie RS, Runcie MJ. Household infrastructure in Aboriginal communities and the implications for health improvement. Med J Aust 2001; 175(7):363–366.

19. Australian Institute of Health and Welfare (AIHW). Housing circumstances of First Nations people. AIHW; 2023 [cited Dec, 2024]. Available from: https://www.aihw.gov.au/reports/australias-welfare/indigenous-housing

20. Australian Building Codes Board (ABCB). National Construction Code (NCC) 2022, Volume 2 - Building Code of Australia Class 1 and 10 Buildings, Part H4: Health and amenity. ABCB; 2022 [cited August, 2025]. Available from: https://ncc.abcb.gov.au/editions/ncc-2022/adopted/volume-two/h-class-1-and-10-buildings/part-h4-health-and-amenity

21. Northern Territory Government (NT Gov). Looking after your public housing home. NT Gov Department of Housing, Local Government, and Community Development; n. d. [cited Dec, 2024]. Available from: https://nt.gov.au/property/social-housing/looking-after-your-home/look-after-your-public-housing-home/looking-after-your-laundry?curr=179521&SQ_PAINT_LAYOUT_NAME=multi&print=yes

22. Grealy L, Lea T. Washing and white goods. Catalyst: Feminism, Theory, Technoscience; 2023. [cited Nov, 2024]. Available from: https://catalystjournal.org/index.php/catalyst/article/view/38168

23. Lloyd CR. Washing machine usage in remote Aboriginal communities. Aust N Z J Public Health 1998; 22(6):695–699.

24. Riley B, Klerck M, Markham F et al. The prepay “poverty premium”: perspective on Australia’s Northern Territory prepayment tariff. Energy Res Soc Sci 2025; 127:104189.

25. Anda M, Mathew K, Ho G. Communal ablutions facility for Aboriginal outstations. In: Mansell D, Stewart D. and Walker B. (eds), Technology for Community Development in Australia, South East Asia and the Pacific. Alice Springs, NT: University of Melbourne and Alice Springs College of TAFE; 1990. pp. 8

26. Meiklejohn J. The story behind launching 14 remote laundry services. Brisbane: Orange Sky; 2004 [cited Nov, 2024]. Available from: https://orangesky.org.au/remote-laundry-services/

27. Aboriginal Investment Group. Remote laundries project. 2024. [cited April, 2025]. Available from: https://www.aiggroup.org.au/core-business/remote-laundries/

28. National Heart Foundation of Australia. 2024-25 pre-budget submission: tackling Australia’s biggest killer: heart disease. Canberra: Heart Foundation; 2004. pp. 18

29. Aboriginal Investment Group (AIG). Empowering First Nations communities to end rheumatic heart disease. AIG; n. d. [cited Nov, 2024]. Available from: https://www.remotelaundries.org.au/empowering-first-nations-communities-to-end-rheumatic-heart-disease/

30. Queensland Health. Ending rheumatic heart disease: Queensland First Nations strategy 2021–2024 final report. Brisbane/Meanjin: Queensland Government; 2024. pp. 17 [cited Nov, 2024]. Available: https://www.health.qld.gov.au/_data/assets/pdf_file/0032/1364936/Final-report_EndingRHD_QLD-FN-Strategy2021-2024.pdf

31. KPMG and Aboriginal Investment Group (AIG). A cost benefit analysis of the Remote Laundries Project. AIG; 2021 [cited Feb 2025]. Available from: https://www.remotelaundries.org.au/wp-content/uploads/2021/03/Remote-Laundries-Project-Cost-benefit-Analysis.pdf

32. Canuto KJ, Street C, Smith JA et al. Social impact framework: AIG remote laundries project. Bedford Park, SA: Rural and Remote Health SA and NT, Flinders University; 2023. pp. 24

33. Social Ventures Australia (SVA). Orange Sky Australia: the mobile laundry service. SVA; 2024 [cited Dec, 2024]. Available from: https://www.socialventures.org.au/our-impact/orange-sky-australia-the-mobile-laundry-service/

34. McPhail-Bell K, Bond C, Brough M, Fredericks B. ‘We don’t tell people what to do’: ethical practice and Indigenous health promotion. Health Promot J Aust 2015; 26(3):195–199.

35. Albanese A, McCarthy M. Albanese Labor Government building on investments to Close the Gap. Department of the Prime Minister and Cabinet; 2025 [cited Sept, 2025]. Available from: https://www.pm.gov.au/media/albanese-labor-government-building-investments-close-gap

36. Heart Foundation. Clean sheets, healthy hearts: Aboriginal Investment Group and Heart Foundation thrilled with Remote Laundries announcement. National Heart Foundation of Australia; 2025 [cited July, 2025]. Available from: https://www.heartfoundation.org.au/media-releases/clean-sheets-healthy-hearts

37. Tricco AC, Lillie E, Zarin W et al. PRISMA Extension for Scoping Reviews (PRISMA-ScR): Checklist and Explanation. Annals Internal Med 2018; 169(7):467–473.

38. National Indigenous Australians Agency (NIAA). Community finder. Australian Government NIAA; 2024. [cited Nov, 2024]. Available from: https://www.indigenous.gov.au/community

39. Australian Government Department of Health and Aged Care (DHAC). Modified Monash Model. DHAC; 2024 [cited Nov, 2024]. Available from: https://www.health.gov.au/topics/rural-health-workforce/classifications/mmm

40. Huria T, Palmer SC, Pitama S et al. Consolidated criteria for strengthening reporting of health research involving Indigenous peoples: the CONSIDER statement. BMC Med Res Methodol 2019; 19(1):173.

41. Chlanda E. Trachoma campaign in Centre part of ending Australia’s shame. Alice Springs News; 2019 [cited Nov, 2024]. Available from: https://alicespringsnews.com.au/2019/03/15/trachoma-campaign-in-centre-part-of-ending-australias-shame/

42. Luke J, Verbunt E, Zhang A et al. Questioning the ethics of evidence-based practice for Indigenous health and social settings in Australia. BMJ Glob Health 2022; 7(6).

43. Australian Institute of Health and Welfare (AIHW). Changing patterns of mortality in Australia since 1900; 2022. pp. 44 [cited Nov, 2024]. Available: https://www.aihw.gov.au/getmedia/2f534d2b-8bf7-42c7-9192-9a328559765d/aihw-aus-240_chapter_4.pdf.aspx

44. Larkin S. Evidence-based policy making in Aboriginal and Torres Strait Islander health. Aust Aboriginal Studies 2006; (2):17–26.

45. Naylor RA, Boase CJ. Practical solutions for treating laundry infested with Cimex lectularius (Hemiptera: Cimicidae). J Economic Entomol 2010; 103(1):136–139.

46. Izri A, Chosidow O. Efficacy of machine laundering to eradicate head lice: recommendations to decontaminate washable clothes, linens, and fomites. Clin Infect Dis 2006; 42(2):e9–10.

47. Akhoundi M, Nasrallah J, Marteau A et al. Effect of household laundering, heat drying, and freezing on the survival of dermatophyte conidia. J Fungi 2022; 8(5):546.

48. Riley K, Williams J, Owen L et al. The effect of low‐temperature laundering and detergents on the survival of Escherichia coli and Staphylococcus aureus on textiles used in healthcare uniforms. J Appl Microbiol 2017; 123(1):280–286.

49. Bockmühl DP, Schages J, Rehberg L. Laundry and textile hygiene in healthcare and beyond. Microbial Cell 2019; 6(7):299.

50. Standards Australia. Australian Standard AS 4146: 2024: Laundry practice. Standards Australia; 2024. pp. 47 [cited Dec, 2024]. Available: https://www.standards.org.au/standards-catalogue/standard-details?designation=as-4146-2024

51. Whitehead K, Eppinger J, Srinivasan V et al. Potential for microbial cross contamination of laundry from public washing machines. Microbiol Res 2022; 13(4):995–1006.

52. Callewaert C, Van Nevel S, Kerckhof F-M et al. Bacterial exchange in household washing machines. Front Microbiol 2015; 6:1381.

53. Summer K, Daw J, Burgess R et al. Fabric contamination and effective laundering for managing skin conditions: a systematic review. Prospero registration: CRD42024594116; 2025.

54. Remote Pools Project. The why. Remote Pools Project and YMCA; n. d. [cited Dec, 2024]. Available from: https://www.remotepoolsproject.ymca.org.au/the-why

55. Scarr J, Roberts F. Remote pools: a Royal Life Saving review of swimming pools in remote areas of the Northern Territory. Darwin, NT: Royal Life Saving; 2010. pp. 20

56. Royal Life Saving WA. More swimming pools needed in remote Aboriginal communities. 2016. [cited Dec, 2024]. Available from: https://royallifesavingwa.com.au/news/community/more-swimming-pools-needed-in-remote-aboriginal-communities

57. Hendrickx D, Stephen A, Lehmann D et al. A systematic review of the evidence that swimming pools improve health and wellbeing in remote Aboriginal communities in Australia. Aust N Z J Public Health 2016; 40(1):30–36.

58. Sanchez L, Carney S, Estermann A et al. An evaluation of the benefits of swimming pools for the hearing and ear health of young Indigenous Australians: a whole of population study across multiple remote Indigenous communities. Adelaide, SA: Flinders University; 2012. pp. 81

59. END RHD Centre of Research Excellence. The RHD Endgame Strategy: Evidence Brief #1. Increasing access to swimming pools and water parks in remote communities. Perth, WA: Telethon Kids Institute; 2020. pp. 2

60. Hall G, Sibthorpe B. Health benefits of swimming pools in remote Aboriginal communities. BMJ 2003; 327(7412):407–408.

61. National Aboriginal Community Controlled Health Organisation (NACCHO). NACCHO policy position paper: Aboriginal housing for Aboriginal health. NACCHO; 2021 [cited Dec, 2024]. Available from: https://www.naccho.org.au/naccho-policy-position-paper-aboriginal-housing-for-aboriginal-health

62. Wyber R, Summer K, Stacey I et al. The need for community-controlled tools to monitor health impacts of housing improvement initiatives in Australia. Front Public Health 2025; 13:1–6.

