## Supplementary File for "The growth of rural and remote Aboriginal and Torres Strait Islander community laundries: an integrative scoping review"

---

### SEARCH STRATEGY

**Table S1:** Terms used to search scientific databases for published literature relevant to the establishment, operation or evaluation of rural and remote Aboriginal or Torres Strait Islander community laundries in Australia since the year 2000 (as of November 2024).

| | |
|---|---|
| 1 | Aboriginal OR Torres Strait Island* OR Indigenous OR “remote community” |
| 2 | Laundry OR laund* OR “washing machine” OR “healthy living practice 2” OR “washing cloth* and bedding” OR “orange sky” OR “Aboriginal investment group” OR AIG |
| 1 AND 2 | All fields, English, 2000-2024 |
| Scopus | 1 hit, 0 relevant |
| Pubmed | 4 hits, 0 relevant |
| Informit Indigenous health collection | 1 hit, 0 relevant |
| Total | 6 hits, 0 duplicates, 0 relevant |

### CONDUCT AND POSITIONALITY

**Table S2:** The ways by which each domain of the CONSIDER statement [1] was addressed in this research.

| | |
|---|---|
| **1. GOVERNANCE** | |
| 1.1 Partnership agreements<br>1.2 Accountability and review mechanisms<br>1.3 Protection of Indigenous intellectual property | – This research is underpinned by Standards for the Conduct of Aboriginal Health Research (The Kids Research Institute of Australia) and standards of associated organisations that are consistent with best practice advice from the NHMRC (https://www.nhmrc.gov.au/research-policy/ethics/ethical-guidelines-research-aboriginal-and-torres-strait-islander-peoples).<br>– It forms part of the NHMRC-funded Stopping Acute Rheumatic Fever to Strength Health (STARFISH) program of work. The hallmark of the STARFISH program is the agenda-setting and leadership of an Indigenous Governance Council (IGC). The rationale, methodology, aims and implications of this research were co-developed by lead investigators and the IGC. These mechanisms emphasise the importance of Indigenous governance, ensure accountability and address harm minimisation.<br>– RB and BJ are members of the STARFISH IGC and have contributed to all phases of this research.<br>– The research supports Aboriginal and Torres Strait Islander self-determination in community laundry service decision making and delivery.<br>– Specific community names or locations have not been reported or discussed in the manuscript. |
| **2. PRIORITISATION** | |
| 2.1 Prioritisation of research aims | – The rationale for this research emerged from the priorities of researchers, clinicians, industry stakeholders, and community partners, many of whom are engaged in work to address skin infections, ARF and RHD; the rollout of community laundries promising health and wellbeing benefits is of interest to all stakeholders, both Indigenous and non-Indigenous.<br>– This research is agnostic to the laundry service provider or model of service delivery prioritising a focus on access to HLP2. |
| **3. RELATIONSHIPS** | |
| 3.1 Measures that adhere to and honour Indigenous ethical guidelines, processes and approvals<br>3.2 Involvement of Indigenous stakeholders in the research<br>3.3 Expertise of the research team | – Ongoing engagement and collaboration with the IGC, as well as informal discussions with wider Indigenous colleagues, community members, laundry service providers, and health care providers demonstrate the commitment of the research team to relationships, and respect for Indigenous stakeholders, culture and histories.<br>– We seek to foster constructive discussion and develop collaborations between stakeholders in what is currently a politicised and potentially divisive space.<br>– Ethics approvals were not required to undertake this work.<br>– RW is a non-Indigenous general practitioner experienced in research and clinical practice in remote Indigenous community settings; she is the lead author of the RHD Endgame Strategy and works as a senior research fellow with The Kids and Yardhura Walani- the National Centre for Aboriginal and Torres Strait Islander Wellbeing Research at the Australian National University.<br>– RB is a Wonnarua woman currently residing in Kununurra. She is an experienced social scientist and senior research fellow, specialising in culturally safe health promotion, and is the Indigenous lead of the END RHD Program at The Kids.<br>– BJ is a Murrawarri man and clinician currently residing in Darwin. He is a Rhodes scholar with the University of Oxford and is undertaking a PhD focused on RHD screening in remote communities.<br>– KS, JD and DN are non-Indigenous early career researchers working in Aboriginal and Torres Strait Islander environmental health. |
| **4. METHODOLOGIES** | |
| 4.1 Rationale of the methods and | – In the absence of scientific literature, we adopted a structured approach to searching for publicly available information; inclusion criteria were thoroughly discussed between co-authors and were strictly context dependent. |

| | |
|---|---|
| implications for Indigenous stakeholders<br>4.2 Consideration of the physical, social, economic and cultural environment of participants | – Both quantitative and qualitative data and methods of evaluation were considered.<br>– We acknowledge that the impacts of colonisation, racism and social injustice are deep and ongoing. It is of utmost importance that community laundry services do not unintentionally contribute to these impacts. The manuscript discusses the importance of cultural safety in remote community laundry service delivery and evidence-based public health promotion of specific health impacts.<br>– We acknowledge the contextual realities precluding access to HLP2 in some remote communities, and the barriers that may be experienced by individuals and service providers. This research will inform evolving pieces of work with specific aims to support effective HLP2, such as a systematic review of effective laundry processes targeting skin pathogens/ectoparasites and the development of technical guidance for HLP2. |
| **5. PARTICIPATION** | |
| 5.1 Consent to conduct future analysis on samples<br>5.2 Resource demands placed on Indigenous participants and communities<br>5.3 Storage and disposal of biological samples | – There were no samples collected, or Indigenous research participants involved in this research.<br>– The recommendation to develop substantiating evidence surrounding remote community laundries will necessitate place-based research. This should not be burdensome to communities nor without meaningful service. Data should be collected, controlled and communicated by Indigenous stakeholders and communities as far as possible. RB and BB, along with the IGC and community partners will continue to inform this work. |
| **6. CAPACITY** | |
| 6.1 Indigenous research capacity<br>6.2 Research team professional development | – This research has inspired discussion and advanced the knowledge of our research team, including Indigenous co-authors, and Indigenous stakeholders, regarding the health and wellbeing impact of HLP capacity building initiatives. It will support new and ongoing employment opportunities for Aboriginal and Torres Strait Islander researchers as work to evaluate the impact of community laundries develops.<br>– All authors seek regular professional development opportunities in Indigenous health and research.<br>– Engagement with the IGC continues to result in two-ways knowledge sharing and benefits for both Indigenous and non-Indigenous authors and collaborators. |
| **7. ANALYSIS AND INTERPRETATION** | |
| 7.1 How the analysis and reporting supported critical enquiry and a strength-based approach | – Concerted effort was made to avoid the use of deficit language surrounding disease burden and living conditions, rather focusing on framing as strengths-based capacity building for HLP2. Specific review of framing and wording was sought from BJ and RB.<br>– BJ and RB contributed to the interpretation and reporting of data, and manuscript review and editing. |
| **8. DISSEMINATION** | |
| 8.1 Dissemination of findings to Indigenous governing bodies and peoples<br>8.2 Process for knowledge translation and implementation to support Indigenous advancement | – End users of this work are wide ranging and expected to include Aboriginal and Torres Strait Islander community organisations, laundry service providers, policymakers, health promotion practitioners, clinicians, and researchers. We will exchange findings with key stakeholders at a workshop and in personal communications.<br>– The results of this review and companion pieces will be presented to Aboriginal and Torres Strait Islander governing bodies at relevant conferences and meetings.<br>– The manuscript acknowledges the need for evaluation of skin infections, ARF and RHD to inform ethical health promotion and support decision making by communities. |

### DATA SUMMARY

**Table S3.** Summary of data collected from the scoping review of community laundries located throughout rural and remote Australia recently established (2000-2024) with the aim of increasing access to HLP2 among Aboriginal and Torres Strait Islander people.

| Comm-unity ID | Juris-diction (region)[1] | MMM [2] | Community pop. size range[3] | Year laundry opened | Provider category[4] | Infrastructure type | Laundry / site count | Facilities[5] | Available financial data | Publicly available notes on health and wellbeing aims and impact | Refs |
|---|---|---|---|---|---|---|---|---|---|---|---|
| Community 1 | NT (Central Australia) | 7 | 201-500 | 2012 | Initiated by ACCO(s) with support from Gov and/or Industry | Permanent laundry structure (improved existing structure) | 1 | 2 WM, 2 D | NA | • "[Washing] will make a big difference to the community and make people more comfortable."<br>• "[It will] offer only a few extra employment opportunities." | 2 |
| Community 2 | NT (Central Australia) | 7 | 201-500 | 2014 | Initiated by Gov, operated in partnership with ACCO(s) | Permanent laundry structure (improved existing structure) | 1 | NA | NA | • Key objective to reduce scabies and skin infections (as part of broader healthy skin program)<br>• "Health improvements in trachoma incidence are strongly suggestive of improved conditions of hygiene. "<br>• Outcomes included greater understanding of skin sores and hygiene | 3-5 |
| Community 3 | NT (Central Australia) | 7 | 25-100 | 2016 | Initiated by LSP, operated in partnership with ACCO(s) | Permanent laundry structure (improved existing structure) | 1 | 1 WM, 1 tub, 1 clothes line | NA | • A functional asset to the community and anticipated to be a new social space | 6,7 |
| Community 4 | NT (Arnhem Land and Groote Eylandt) | 7 | 501-1000 | 2021 | Initiated by LSP, operated in partnership with ACCO(s) | Permanent laundry structure (new structure) | 1 | 4 WM, 4 D, hot water, "top quality detergent" supplied by LSP | NA | • "In the communities where our laundries now operate, they have improved health, improved quality of life [and] created employment for community members."<br>• "A reduction in the incidence of primary and secondary medical conditions associated with bacterial infections."<br>• "Improved quality of life associated with reduced | 8,9 |

| | | | | | | | | | | | |
|---|---|---|---|---|---|---|---|---|---|---|---|
| | | | | | | | | | | • disability and additional medical complications.”<br>• “Social benefits from the direct employment of local staff.” | |
| Community 5 | Qld (North Qld) | 7 | 501-1000 | 2021 | Initiated by LSP, operated in partnership with ACCO(s) | Mobile laundry vehicle, fixed location/s | 1 | 3 WM, 3 D | Free service | • Facilitates local employment and conversations<br>• Supports skin health and overall quality of life<br>• Also provides a shower service<br>• “[Improves] access to health hardware”<br>• “High hygienic wash standards.” | 10-12 |
| Community 6 | NT (Top End and Tiwi Islands) | 7 | 201-500 | 2019 | Initiated by LSP, operated in partnership with ACCO(s) | Permanent laundry structure (new structure) | 1 | 4 WM, 4 D | $224,660 in year 1, ~$110,000 annual thereafter; initially cost $4 to WM and $4 to use D but now free to increase access | • 60% drop in scabies cases over 4 years<br>• Strong links between scabies and post-streptococcal complications<br>• "This will help deal with scabies and trachoma, the laundry will help solve all these problems when people can wash their clothes better."<br>• “More laundries mean less scabies.”<br>• “Washing clothes, bedding, and towels in hot water with medical grade detergent kills the scabies mite, and if done often enough will reduce mite populations within any household.”<br>• “We need to be acting now to protect the health of all people living in communities. We believe our laundries are a valuable weapon in the fight.”<br>• “With enough laundries throughout the NT we can bring down unacceptable incidence of scabies, rheumatic heart disease and kidney disease.” | 9,13-17 |
| Community 7 | WA (Kimberley) | 7 | 501-1000 | 2021 | Initiated by LSP, operated in partnership with ACCO(s) | Mobile laundry trailer, fixed location/s | 1 | 3 WM, 3 D | NA | | 18 |

| | | | | | | | | | | | |
|---|---|---|---|---|---|---|---|---|---|---|---|
| Community 8 | Qld (North Qld) | 7 | 1001-1500 | 2023 | Initiated by LSP, operated in partnership with ACCO(s) | Mobile laundry vehicle, fixed location/s | 1 | 3 WM, 3 D | NA | • As part of "commitment to ending RHD by 2031"<br>• "Clean clothing is essential to reducing the risks of skin infections… the mobile laundry is a critical step towards ensuring access to essential hygiene infrastructure." | 19,20 |
| Community 9 | WA (Kimberley) | 7 | 1001-1500 | 2021 | Initiated by LSP, operated in partnership with ACCO(s) | Mobile laundry vehicle, temporary service | 1 | 3 WM, 3 D | Free service | • [Lack of access] has consequences for physical, social and emotional wellbeing. | 10,21 |
| Community 10 | NT (Arnhem Land and Groote Eylandt) | 7 | 2001-2050 | 2022 | Initiated by LSP, operated in partnership with ACCO(s) | Mobile laundry vehicle, temporary service | 1 | 3 WM, 3 D | Free service | | 10,22 |
| Community 11 | NT (Arnhem Land and Groote Eylandt) | 7 | 501-1000 | 2023 | Initiated by LSP, operated in partnership with ACCO(s) | Mobile laundry vehicle, fixed location/s | 1 | 3 WM, 3 D | Free service | • "[The laundry] will provide a way for residents to ensure good skin health and prevent the development of chronic disease (ARF/RHD)."<br>• "Clean bedding is vital in promoting good health.. [it] is crucial for preventing the spread of scabies and other skin infections."<br>• Improves overall health, sleep, and well-being. | 23 |
| Community 12 | NT (Arnhem Land and Groote Eylandt) | 7 | 201-500 | 2023 | Initiated by LSP, operated in partnership with ACCO(s) | Mobile laundry vehicle, fixed location/s | 1 | 3 WM, 3 D | NA | | 24 |
| Community 13 | NT (Arnhem Land and Groote Eylandt) | 7 | 1001-1500 | 2024 | Initiated by LSP, operated in partnership with ACCO(s) | Permanent laundry structure (new structure) | 1 | 4 WM, 4 D | The remote laundry program has so far injected over $650,000 of wages into remote | • "Provides free washing and drying for the community to reduce the prevalence of scabies and other skin infections and improve health and wellbeing."<br>• "[A] trusted solution to combat the burden of disease in remote communities while creating jobs." | 25,26 |

| | | | | | | | | | | | |
|---|---|---|---|---|---|---|---|---|---|---|---|
| | | | | | | | | | communities | | |
| Community 14 | WA (Greater WA) | 3 | 2001-2500 | 2023 | Initiated by LSP, operated in partnership with ACCO(s) | Mobile laundry vehicle, fixed location/s | 2 | 3 WM, 3 D | Free service | • "Will allow our social support staff to make contact with people who don't normally come into the service and perhaps assist them to get help with the issues contributing to their circumstance"<br>• "A means to achieving meaningful outcomes for people who are homeless or experiencing hardship." | 24,27,28 |
| Community 15 | WA (Kimberley) | 7 | 201-500 | 2017 | Initiated by ACCO(s) with support from Gov and/or Industry | Permanent laundry structure (new) | 1 | NA | NA | • Commissioned in response to the APSGN outbreak in 2014-2015 | 29 |
| Community 16 | Qld (North Qld) | 7 | 501-1000 | 2024 | Initiated by LSP, operated in partnership with ACCO(s) | Mobile laundry trailer, fixed location/s | 1 | 3 WM, 3 D | Free service | | 30 |
| Community 17 | WA (Kimberley) | 7 | 1001-1500 | 2024 | Initiated by LSP, operated in partnership with ACCO(s) | Mobile laundry vehicle, fixed location/s | 5 | 3 WM, 3 D | Free service | • "Our laundry facility has a water heater to make sure every wash is a hot wash. A 30-minute wash with hot water can kill harmful bacteria and kill scabies mites and we know this is important for skin health. A hot water wash is also important when washing after natural disasters to kill harmful bacteria."<br>• "The washing detergent is specially formulated.. it includes an antibacterial agent and sanitiser, and doesn't contain fragrance, so it's suitable for people with sensitive skin." | 24,31 |
| Community 18 | NT (Top End and Tiwi Islands) | 7 | 501-1000 | 2023 | Initiated by ACCO(s) with support from Gov and/or Industry | Permanent laundry structure (new structure) | 1 | NA | $406,863 to design and build | | 32 |
| Community 19 | Qld (North Qld) | 7 | 201-500 | 2017 | Initiated by LSP, operated | Temporary fixed laundry | 1 | NA | NA | • Forms part of Qld Ending RHD Strategy 2021-2024 | 33,34 |

| | | | | | | | | | | | |
|---|---|---|---|---|---|---|---|---|---|---|---|
| | | | | | in partnership with ACCO(s) | structure (pod) (new) | | | | | |
| Community 20 | NT (Central Australia) | 7 | 501-1000 | 2019 | Initiated by LSP, operated in partnership with ACCO(s) | Mobile laundry trailer, fixed location/s | 1 | 1 WM (?) (manual) | Cost $14, 000 to install and fit out | • "We work with communities, to identify what their capacity allows and how they would like to improve hygiene and sanitation in their communities."<br>• "With every kid's face clean, trachoma would disappear by the end of the year." | 35 |
| Community 21 | NT (Arnhem Land and Groote Eylandt) | 7 | 2001-2500 | 2020 | Initiated by LSP, operated in partnership with ACCO(s) | Temporary fixed community laundry structure (pod);<br><br>Mobile laundry vehicle, fixed location/s | 1<br><br>1 | 2 WM, 2 D<br><br>3 WM, 3 D | Free service | • "[The LSP} shared our vision to improve health outcomes for people in remote communities. In a community with high numbers of RHD, a partnership with [the LSP] would make a significant difference to the health of our people." | 36,37 |
| Community 22 | NT (Arnhem Land and Groote Eylandt) | 7 | 101-200 | 2022 | Initiated by LSP, operated in partnership with ACCO(s) | Permanent laundry structure (new structure) | 1 | 4 WM, 4 D | NA | • "Our laundries not only address the health issues associated with a lack of washing facilities in remote communities; they create employment opportunities and a hub for conversation and education."<br>• "Through workshops, staff were able to build their knowledge around the environmental factors that contribute to ARF and RHD, such as skin sores and scabies and the preventative actions and protections that prevent the spread."<br>• "Promoting clean and healthy living, assisting in the prevention of skin sores and providing local employment is a key necessity to advancing health." | 38-40 |
| Community 23 | SA (APY lands) | 7 | 201-500 | 2013 | Initiated by Gov, operated | Permanent laundry | 1 | 4 WM, undercover clothes | NA | | 41 |

| | | | | | | | | | | | |
|---|---|---|---|---|---|---|---|---|---|---|---|
| | | | | | in partnership with ACCO(s) | structure (new structure) | | lines, tokens and detergent sachets purchased from the store | | | |
| Community 24 | NT (Central Australia) | 7 | 201-500 | 2017 | Initiated/ operated by ACCO(s) | Permanent laundry structure (improved existing structure) | 1 | NA | NA | | 42,43 |
| Community 25 | NT (Central Australia) | 7 | 201-500 | 2018 | Initiated by ACCO(s) with support from Gov | Permanent laundry structure (new structure) | 1 | 7 WM/D, sinks and bench space | NA | • "This initiative has been welcomed by the community, as it provides a place for washing clothes and larger items such as blankets, in turn improving hygiene and health outcomes." | 44 |
| Community 26 | NT (Top End and Tiwi Islands) | 7 | 201-500 | 2020 | Initiated by LSP, operated in partnership with ACCO(s) | Temporary fixed community laundry structure (pod) | 1 | 2 WM, 2 D | Free service | • "Cultural integrity and independence at the forefront to improve wellbeing and outcomes in the communities where we operate." | 12,45 |
| Community 27 | Qld (North Qld) | 7 | 3501-4000 | 2024 | Initiated by LSP, operated in partnership with ACCO(s) | Mobile laundry vehicle, fixed locations | 2[6] | 3 WM, 3 D | Free service | • Was initiated "in response to increased skin health and wellbeing concerns." | 24,46 |
| Community 28 | NT (Central Australia) | 7 | 25-100 | 2020 | Initiated by Gov, operated in partnership with ACCO(s) | Permanent laundry structure (improved existing structure) | 1 | 1 WM | $74,000 to install | | 47 |
| Community 29 | Qld (North Qld) | 7 | 1501-2000 | 2018 | Initiated by LSP, operated in partnership with ACCO(s) | Mobile laundry vehicle, fixed location/s | 8 | 3 WM, 3 D | Free service | • "These vehicles are solar powered and deliver hot washing to improve hygiene."<br>• "Listening and learning to ensure our service aligns with the community's health, social and cultural needs." | 12,24,48 |
| Community 30 | WA (Greater WA) | 7 | 25-100 | 2023 | Initiated by Gov, operated in partnership with ACCO(s) | Permanent laundry structure (improved | 1 | NA | Part of a $1.17 million grant for | • "Helping to develop resilient and strong Aboriginal communities." | 49 |

| | | | | | | | | | | | |
|---|---|---|---|---|---|---|---|---|---|---|---|
| | | | | | | existing structure) | | | infrastructure and service upgrades | | |
| Community 31 | NT (Central Australia) | 7 | 25-100 | 2007 | Initiated by ACCO(s) with support from Gov and/or Industry | Permanent laundry structure (improved existing structure) | 1 | NA | NA | | 50 |
| Community 32 | NT (Top End and Tiwi Islands) | 7 | 1501-2000 | 2021, 2023 | Initiated by LSP, operated in partnership with ACCO(s) | Mobile laundry vehicle, fixed location/s | 2 | 6 WM, 6 D | Free service | • “This service aims to improve social and health outcomes for the community.”<br>• “Washing is not merely a chore and is much more than a hygiene solution. It plays an important role in people’s social and physical well-being. It also provides employment and training opportunities for local people.” | 24,51,52 |
| Community 33 | WA (Greater WA) | 7 | 25-100 | 2009 | Initiated by ACCO(s) with support from Gov and/or Industry | Permanent laundry structure (improved existing structure) | 1 | 3 WM, 2 D, basin | Tokens cost $4 for 1 WM or D load | | 53 |
| Community 34 | WA (Kimberley) | 7 | 101-200 | 2022 | Initiated by LSP, operated in partnership with ACCO(s) | Mobile laundry vehicle, temporary service | 1 | 3 WM, 3 D | Free service | | 10,22 |
| Community 35 | Qld (North Qld) | 5 | 1001-1500 | 2019 | Initiated by ACCO(s) with support from State/Territory government | Permanent laundry structure (improved existing structure) | 1 | NA | NA | | 54 |
| | | | | 2019 | Initiated by LSP, operated in partnership with ACCO(s) | Mobile laundry vehicle, temporary service | 1 | 3 WM, 3 D | Free service | | 55 |
| Community 36 | NT (Central Australia) | 7 | 1501-2000 | 2019 | Initiated by ACCO(s) with support from Gov and/or Industry | Permanent laundry structure (improved | 1 | NA | NA | | 56 |

| | | | | | | | | | | | |
|---|---|---|---|---|---|---|---|---|---|---|---|
| | | | | | | existing structure) | | | | | |
| | | | | 2023 | Initiated by LSP, operated in partnership with ACCO(s) | Mobile laundry vehicle, temporary service | 1 | 3 WM, 3 D | Free service | | 12,57 |
| Community 37 | WA (Kimberley) | 7 | 201-500 | 2022 | Initiated by LSP, operated in partnership with ACCO(s) | Mobile laundry vehicle, temporary service | 1 | 3 WM, 3 D | Free service | | 10,12 |
| Community 38 | NT (Central Australia) | 7 | 501-1000 | 2024 | Initiated by LSP, operated in partnership with ACCO(s) | Permanent laundry structure (improved existing structure) | 1 | 3 WM | | | 58 |

[1.] Jurisdictions: NT: Northern Territory, WA: Western Australia, Qld: Queensland, SA: South Australia; regions correspond to the NIAA map (Figure S1)

[2.] Remoteness classification according to the Modified Monash Model [59]

[3.] Aboriginal or Torres Strait Islander community population size was sourced from Australian Bureau of Statistics census data 60 or the NT Government BushTel website 61. Ranges were pre-defined by the authors in increments of 100, 200 or 500. These figures are intended to be approximate only for the purpose of comparing the ratio of laundry facilities to people

[4.] LSP: laundry service provider, where major providers are Orange Sky and Aboriginal Investment Group [AIG]; ACCO: local (land council level) Aboriginal or Torres Strait Islander community-controlled organisation; Gov refers to State/Territory Government department/organisation

[5.] WM: washing machines, D: dryers, and consumables

[6.] Of five fixed locations in the town, two are specifically intended to increase access for Aboriginal people

NA: data not available

### References

1. Huria T, Palmer SC, Pitama S *et al*. Consolidated criteria for strengthening reporting of health research involving Indigenous peoples: the CONSIDER statement. *BMC Med Res Methodol* 2019; 19(1):173.
2. Brain, C. Art and laundry expands in Ali Curung. ABC News; 2012 [cited Nov, 2024]. Available from: https://www.abc.net.au/news/rural/2012-07-25/art-and-laundry-expands-in-ali-curung/6159932
3. Barkly Regional Council. Communities: Alpurrurulam and LA meetings. n. d. [cited Nov, 2024]. Available from: https://www.barkly.nt.gov.au/communities/alpurrurulam
4. Human Capital Alliance (HCA). Evaluation snapshot: Alpurrurulam community wellness and Utopia community gardens. Northern Territory Medicare Local and HCA; 2014 [cited Nov, 2024]. Available from: https://humancapitalalliance.com.au/wp-content/uploads/2015/12/2015-03-Evaluation-Snapshot_FINAL_print.pdf
5. Thomas K. Lessons learnt from capacity building community projects in remote Central Australia. Paper presented at the 13th National Rural Health Conference, Darwin NT, May 2015. pp. 4 [cited Nov, 2024]. Available from: https://www.ruralhealth.org.au/13nrhc/images/paper_Thomas,%20Karen.pdf
6. Central Land Council. Community Development News, Summer 2016: In the Wash in Alyuen. 2016. pp. 9 [cited Nov, 2024]. Available from: https://www.clc.org.au/wp-content/uploads/2021/03/CLC-Community-Development-News-Summer-2016.pdf
7. Centre for Appropriate Technology. Alyuen laundry. n. d. [cited Nov, 2024]. Available from: https://www.cfat.org.au/alyuen-laundry
8. Aboriginal Investment Group (AIG). Angurugu. n. d. [cited Nov, 2024]. Available from: https://www.remotelaundries.org.au/angurugu/
9. KPMG and Aboriginal Investment Group (AIG). A cost benefit analysis of the Remote Laundries Project. AIG; 2021 [cited Feb 2025]. Available from: https://www.remotelaundries.org.au/wp-content/uploads/2021/03/Remote-Laundries-Project-Cost-benefit-Analysis.pdf
10. Meiklejohn J. The story behind launching 14 remote laundry services. Brisbane: Orange Sky; 2004 [cited Nov, 2024]. Available from: https://orangesky.org.au/remote-laundry-services/
11. Orange Sky. Aurukun. 2024 [cited Nov, 2024]. Available from: https://orangesky.org.au/locations/aurukun/
12. Orange Sky. Remote communities. 2025 [cited April, 2025]. Available from: https://orangesky.org.au/remote-communities/
13. Aboriginal Investment Group. Barunga. n. d. [cited Nov, 2024]. Available from: https://www.remotelaundries.org.au/barunga/
14. Aboriginal Investment Group. Proof our remote laundries project is reducing scabies. 2021 [cited Nov, 2024]. Available from: https://www.remotelaundries.org.au/proof-our-remote-laundries-project-is-reducing-scabies/
15. Bardon, J. Clean break for Barunga as new laundry opens up to keep scabies at bay. ABC News; 2019 [cited Nov, 2024]. Available from: https://www.abc.net.au/news/2019-03-02/laundry-scabies-remote-community-northern-land-council-health/10861480
16. Aboriginal Investment Group. Is our laundry valuable for the people of Barunga? n. d. [cited Dec, 2024]. Available from: https://www.remotelaundries.org.au/are-public-laundries-aboriginal-communities-valuable/
17. Aboriginal Investment Group. War on scabies: Barunga laundry good news stories. n. d. [cited Dec, 2024]. Available from: https://www.remotelaundries.org.au/war-on-scabies/
18. Orange Sky. Bidyadanga. 2024 [cited Nov, 2024]. Available from: https://orangesky.org.au/locations/bidyadanga/
19. Orange Sky. Doomadgee. 2024 [cited Nov, 2024]. Available from: https://orangesky.org.au/locations/doomadgee/
20. North West Hospital and Health Service. North West HHS partners with Orange Sky and Doomadgee Aboriginal Shire Council to improve health outcomes in Doomadgee. Queensland Government; 2023 [cited Nov, 2024]. Available from: https://www.northwest.health.qld.gov.au/north-west-hhs-partners-with-orange-sky-and-doomadgee-aboriginal-shire-council-to-improve-health-outcomes-in-doomadgee/
21. Orange Sky. Impact Report, 2021/2022. 2022 [cited Nov, 2024]. Available from: https://orangesky.org.au/wp-content/uploads/2024/06/OrangeSky-Impact-Report-2021-2022-Financials_Hires.pdf
22. Orange Sky. Impact Report 2022/2023. 2023 [cited Nov, 2024]. Available from: https://orangesky.org.au/wp-content/uploads/2024/06/AU-OrangeSky-ImpactReport-22-23-with-financials-Condensed.pdf
23. Mitwatj Health Aboriginal Corporation. Orange Sky Laundry in Gapuwiyak. 2023 [cited Nov, 2024]. Available from: https://www.miwatj.com.au/blog/orange-sky-laundry-gapuwiyak/
24. Orange Sky. Our locations. 2024 [cited Nov, 2024]. Available from: https://orangesky.org.au/our-locations/

25. WIRE. New laundry in Gunbalanya. West Arnhem Regional Council; 2024 [cited Nov, 2024]. Available from: https://westarnhem.nt.gov.au/sites/default/files/2024-10/West%20Arnhem%20Wire_515%2028%20September%20-%2011%20October%202024_web_0.pdf
26. Aboriginal Investment Group. West Arnhem community in a spin over new laundry. 2024 [cited Nov, 2024]. Available from: https://www.remotelaundries.org.au/west-arnhem-community-in-a-spin-over-new-laundry/
27. Chounding, A. Kalgoorlie, Boulder Orange Sky Laundry service to support Aboriginal health services. ABC News; 2023 [cited Nov, 2024]. Available from: https://www.abc.net.au/news/2023-02-11/orange-sky-laundry-service-starts-in-kalgoorlie/101942848
28. Orange Sky. Kalgoorlie. 2024 [cited Nov, 2024]. Available from: https://orangesky.org.au/locations/kalgoorlie/
29. McKay S, Prouse I. “You gotta wash ya face to come to my place” community project. 11th National Aboriginal and Torres strait Islander Environmental Health Conference, Cairns, Queensland, 2017. pp. 100
30. Orange Sky. Mornington Island. 2024 [cited Nov, 2024]. Available from: https://orangesky.org.au/locations/mornington-island/
31. Orange Sky. Kununurra. 2024 [cited Nov, 2024]. Available from: https://orangesky.org.au/locations/kununurra/
32. Central Land Council. Central Land Council Annual Report 2022-23: Output 4.4 Community development support. 2023 [cited Nov, 2024]. Available from: https://www.transparency.gov.au/publications/prime-minister-and-cabinet/central-land-council/central-land-council-annual-report-2022-23/output-group-4/output-4.4-community-development-support
33. Orange Sky. Lockhart River. 2024 [cited Nov, 2024]. Available from: https://orangesky.org.au/locations/lockhart-river/
34. Queensland Health. Ending rheumatic heart disease: Queensland First Nations strategy 2021–2024 final report. Brisbane, Queensland: Queensland Government; 2024. pp. 17 [cited Nov, 2024]. Available from: https://www.health.qld.gov.au/__data/assets/pdf_file/0032/1364936/Final-report_EndingRHD_QLD-FN-Strategy2021-2024.pdf
35. Chlanda E. Trachoma campaign in Centre part of ending Australia's shame. Alice Springs, NT: Alice Springs News; 2019 [cited Nov, 2024]. Available from: https://alicespringsnews.com.au/2019/03/15/trachoma-campaign-in-centre-part-of-ending-australias-shame/
36. Orange Sky. Maningrida. 2024 [cited Nov, 2024]. Available from: https://orangesky.org.au/locations/maningrida/
37. Mala'la Health Service Aboriginal Corporation. Celebrating 3-year partnership with Orange Sky Australia. 2023 [cited Nov, 2024]. Available from: https://www.malala.com.au/my-post
38. Aboriginal Investment Group. Milyakburra. n. d. [cited Nov, 2024]. Available from: https://www.remotelaundries.org.au/bickerton-island/
39. Aboriginal Investment Group. Empowering First Nations communities to end rheumatic heart disease. n. d. [cited Nov, 2024]. Available from: https://www.remotelaundries.org.au/empowering-first-nations-communities-to-end-rheumatic-heart-disease/
40. Groote Eyelandt Aboriginal Trust. Charitable purposes: advancing health. 2024 [cited Nov, 2024]. Available from: https://geat.com.au/advancing-health/
41. Government of South Australia, Department of the Premier and Cabinet - Aboriginal Affairs and Reconciliation Division. South Australian Government Update: Progress on the Anangu Pitjantjatjara Yankunytjatjara (APY) Lands. 2013. pp. 37 [cited Nov, 2024]. Available from: https://www.agd.sa.gov.au/__data/assets/file/0003/807780/APY-Lands_Update-June2013.pdf
42. Purple House. Where we work. n. d. [cited Nov, 2024]. Available from: https://www.purplehouse.org.au/communities
43. Remote Area Health Corps. Mount Liebig, Central Region. n. d. [cited Nov, 2024]. Available from: https://www.rahc.com.au/sites/default/files/2022-10/210824_RAHC_Community-Profile_MtLiebig.pdf
44. Australian Government Office of Township Leasing (OTL). Executive Director of Township Leasing Annual Report 2018-19. Darwin, NT: OTL; 2019. pp. 56
45. Orange Sky. Nganmarriyanga. 2024 [cited Nov, 2024]. Available from: https://orangesky.org.au/locations/nganmarriyanga/
46. Orange Sky. Mount Isa. 2024 [cited Nov, 2024]. Available from: https://orangesky.org.au/locations/mount-isa/
47. Central Land Council (CLC). Central Land Council Annual Report 2020–21. Alice Springs, NT: CLC; 2021. pp. 152
48. Orange Sky. Palm Island. 2024 [cited Nov, 2024]. Available from: https://orangesky.org.au/locations/palm-island/
49. Torre, G. Major upgrades planned for Patjarr Community services and infrastructure. National Indigenous Times; 2023 [cited Nov, 2024]. Available from: https://nit.com.au/20-11-2023/8701/major-upgrades-planned-for-patjarr-community-services-and-infrastructure

50. Central Land Council (CLC). Central Land Council Annual Report 2007-2008. Alice Springs, NT: CLC; 2008. pp. 173
51. Orange Sky. Wadeye. 2024 [cited Nov, 2024]. Available from: https://orangesky.org.au/locations/wadeye/
52. Thamarrurr Development Corporation. Washing Warriors TDC Partnership. 2022 [cited Nov, 2024]. Available from: https://thamarrurr.org.au/news/orange-sky-partnership/
53. Shire of Wiluna. Newsletter, Feb 2010, Vol 1 Issue 1. 2010 [cited Nov, 2024]. Available from: https://www.wiluna.wa.gov.au/documents/42/newsletter-february-2010
54. Wellington, S. Indigenous community of Yarrabah opens Youth Hub. SBS/NITV; 2019 [cited Nov, 2024]. Available from: https://www.sbs.com.au/nitv/article/indigenous-community-of-yarrabah-opens-youth-hub/86rwaui3f
55. Yarrabah Aboriginal Shire Council. Yarrabah News. 2019 [cited Nov, 2027]. Available from: http://chowes.com.au/3.19%201119%20Yarrabah%20News.pdf
56. Campbell D, Gyles A. Central Land Council community development program: monitoring report, July 2019-June 2020. Alice Springs, NT: Central Land Council; 2021. pp. 66
57. Orange Sky. Yuendumu. 2024 [cited Nov, 2024]. Available from: https://orangesky.org.au/locations/yuendumu/
58. EndTrachoma. EndTrachoma installs community laundry in Ampilatwatja. 2024 [cited Dec, 2024]. Available from: https://www.endtrachoma2020.org.au/
59. Australian Government Department of Health and Aged Care. Modified Monash Model. 2024 [cited Nov, 2024]. Available from: https://www.health.gov.au/topics/rural-health-workforce/classifications/mmm
60. Australian Bureau of Statistics. Search Census Data. 2024 [cited Available from: https://www.abs.gov.au/census/find-census-data/search-by-area
61. Northern Territory Government. BushTel: Communities A-Z. 2024 [cited Nov, 2024]. Available from: https://bushtel.nt.gov.au/
